## Supplemental Table 1 for "A systematic review of Hepatitis B virus (HBV) prevalence and genotypes in Kenya: Data to inform clinical care and health policy"

**Supplementary Table 2: Joanna Briggs Checklist for Prevalence Studies (Available from https://jbi.global/critical-appraisal-tools).**

| **First Author** | Was the sample frame appropriate to address the target population? | Were the participants sampled in an appropriate way | Was the sample size adequate | Study subjects and setting described in detail? | Was the data analysis conducted with sufficient coverage of the identified sample? | Were valid methods used for the identification of the condition? | Was the condition measured in a standard, reliable way for all participants? | Was there appropriate statistical analysis? | Was the response rate adequate, and if not, was the low response rate managed appropriately? |
|---|---|---|---|---|---|---|---|---|---|
| Ngaira JA (1) | Yes | Yes | Yes | Yes | Yes | Yes ELISA | Yes | Yes | Yes |
| Awili HO (2) | Yes | Yes | No calculation | Yes | Yes | Yes ELISA | Yes | Yes | Yes |
| Onyango CG (3) | Yes | Yes | No calculation | No – looks like recruited from schools | Yes | Yes - ELISA | Yes | Yes | Yes |
| Wamamba D (4) | Yes | Yes | No calculation | Yes | Yes | Yes ELISA | Yes | Yes | Yes |
| Aluora PO (5) | Yes | Yes | Yes | Yes | Yes | Yes – CMIA | Yes | Yes | Mostly young men |
| Bartonjo G (6) | Yes | Yes | ? Intuitive sample size | Yes | Yes | Yes - ELISA | Yes | Yes | Over-represented one area (Nakuru) |
| Kisangau EN (7) | Yes | Yes | Yes | Yes | Yes | Yes - ELISA | Yes | Yes | Mostly female nurses <35 years |
| Ly KN (8) | No – only HIV -ve patients included | Yes | No calculation | Yes | Yes | Yes – ELISA | Yes | Yes | Yes |
| Maina DN (9) | No – only small part of Kenya sampled | Yes | No calculation | Yes | Yes | Yes - CLEIA | Yes | Yes | Yes |

| | | | | | | | | | |
|---|---|---|---|---|---|---|---|---|---|
| Kim HN (10) | No – only in Nairobi | Randomisation technique not described | No calculation | No clear inclusion/exclusion | Yes | Yes – ELISA | Yes | Yes | Yes |
| EN Njuguna (11) | Yes | No – purposive sampling | No calculation | Yes | Yes | Yes – ELISA | Yes | Yes | Yes |
| Muriuki BM (12) | Yes | Yes | No calculation | Yes | Yes | Yes - ELISA | Yes | Yes | Yes |
| Harania RS (13) | No – only done in Nairobi | No description of sampling technique | No calculation | Minimal detail | Yes | Yes – ELISA | Yes | Yes | Yes |
| Greer AE (14) | Yes | Yes | No calculation | No clear inclusion/exclusion | Yes | Yes – CLIA | Yes | Yes | Yes |
| Oyaro M (15) | Yes | Yes | No calculation | Yes | Yes | Yes - ELISA | Yes | Yes | Many more males than females |
| Webale MK (16) | yes | Yes | No calculation | Yes | Yes | Unclear – 5 panel rapid test | Yes | Yes | HIV population over-represented |
| Day SL (17) | Yes | Yes | No calculation | Yes | Yes | Yes – CLIA | Yes | Yes | Yes |
| Ochwoto M (18) | Yes | Yes | No calculation | Yes | Yes | Yes - CLEIA | Yes | Yes | Many more from Nairobi |
| Muchiri I (19) | Yes | No description of sampling | No calculation | No clear inclusion/exclusion | Yes | Reverse passive haemagglutination | Yes | Yes | Yes |
| Atina JO (20) | Yes | Yes | No calculation | Yes | Yes | Reverse passive haemagglutination | Yes | Yes | Yes |
| Wahome E (21) | Yes | Yes | No calculation | Yes | Yes | Yes ELISA | Yes | Yes | High proportion of sex workers |
| Jepkemei (22) | No – only done in Nairobi | Yes | No calculation | Yes | Yes | Yes ELISA and DNA PCR | Yes | Yes | Yes |
| Salyani (23) | Yes | Yes | Yes | Yes | Yes | Yes ELISA and DNA PCR | Yes | Yes | Yes |

1. Malungu Ngaira JA, Kimotho J, Mirigi I, Osman S, Ng'ang'a Z, Lwembe R, et al. Prevalence, awareness and risk factors associated with hepatitis b infection among pregnant women attending the antenatal clinic at mbagathi district hospital in Nairobi, Kenya. Pan African Medical Journal. 2016 Aug 17;24.
2. Awili HO, Gitao GC, Muchemi GM. Seroprevalence and Risk Factors for Hepatitis B Virus Infection in Adolescent Blood Donors within Selected Counties of Western Kenya. BioMed Research International. 2020;2020.
3. Onyango CG, Ogonda L, Guyah B, Okoth P, Shiluli C, Humwa F, et al. Seroprevalence and determinants of transfusion transmissible infections among voluntary blood donors in Homabay, Kisumu and Siaya counties in western Kenya. BMC Research Notes. 2018 Mar 12;11(1).
4. Wamamba D, Onyango D, Oyugi E, Kanyina E, Obonyo M, Githuku J, et al. Transfusion transmissible infections among walk-in blood donors at Kisumu regional blood transfusion centre, Kisumu County, Kenya, 2015. Lab Medicine. 2017 Nov 1;48(4):362–6.
5. Aluora PO, Muturi MW, Gachara G. Seroprevalence and genotypic characterization of HBV among low risk voluntary blood donors in Nairobi, Kenya. Virology Journal. 2020 Dec 1;17(1).
6. Bartonjo G, Oundo J, Ng'ang'a Z. Prevalence and associated risk factors of transfusion transmissible infections among blood donors at regional blood transfusion center nakuru and tenwek mission hospital, Kenya. Pan African Medical Journal. 2019;34.
7. Kisangau EN, Awour A, Juma B, Odhiambo D, Muasya T, Kiio SN, et al. Prevalence of hepatitis B virus infection and uptake of hepatitis B vaccine among healthcare workers, Makueni County, Kenya 2017. Journal of Public Health (United Kingdom). 2019 Dec 1;41(4):765–71.
8. Ly KN, Kim AA, Umuro M, Drobenuic J, Williamson JM, Montgomery JM, et al. Prevalence of hepatitis B Virus infection in Kenya, 2007. American Journal of Tropical Medicine and Hygiene. 2016 Aug 1;95(2):348–53.
9. Maina D, Karen Hospital T, Kimang AN, Mwangi J, Mutai K, Lihana RW. Genotypes of HBV and HCV among HIV-1 co-infected individuals in Ngong Sub-County, Kenya. East African Medical Journal [Internet]. 2017 Mar 30 [cited 2021 Dec 2];93(12):640–5. Available from: https://www.ajol.info/index.php/eamj/article/view/153976
10. Kim HN, Scott J, Cent A, Cook L, Morrow RA, Richardson B, et al. HBV lamivudine resistance among hepatitis B and HIV coinfected patients starting lamivudine, stavudine and nevirapine in Kenya. Journal of Viral Hepatitis. 2011 Oct;18(10).
11. Njuguna EN, Kinuthia PN, Kinoti M, of Public Health S, Kiarie J. Prevalence and risk factors of previous or active Hepatitis B infection among HIV-1 discordant heterosexual couples. East African Medical Journal [Internet]. 2015 Dec 9 [cited 2021 Dec 2];92(10):488–94. Available from: https://www.ajol.info/index.php/eamj/article/view/127082
12. Muriuki BM, Gicheru MM, Wachira D, Nyamache AK, Khamadi SA. Prevalence of hepatitis B and C viral co-infections among HIV-1 infected individuals in Nairobi, Kenya. BMC Research Notes. 2013;6(1).

13. Harania RS, Karuru J, Nelson M, Stebbing J. HIV, hepatitis B and hepatitis C coinfection in Kenya. AIDS. 2008 Jun 19;22(10):1221–2.
14. Greer AE, Ou SS, Wilson E, Piwowar-Manning E, Forman MS, McCauley M, et al. Comparison of hepatitis b virus infection in HIV-infected and HIV-uninfected participants enrolled in a multinational clinical trial: HPTN 052. Journal of Acquired Immune Deficiency Syndromes. 2017;76(4):388–93.
15. Oyaro M, Wylie J, Chen CY, Ondondo RO, Kramvis A. Human immunodeficiency virus infection predictors and genetic diversity of hepatitis B virus and hepatitis C virus co-infections among drug users in three major Kenyan cities. Southern African Journal of HIV Medicine. 2018;19(1).
16. Kilongosi MW, Budambula V, Lihana R, Musumba FO, Nyamache AK, Budambula NLM, et al. Hepatitis B virus sero-profiles and genotypes in HIV-1 infected and uninfected injection and Non-injection drug users from coastal Kenya. BMC Infectious Diseases [Internet]. 2015 Jul 30 [cited 2022 Jan 31];15(1):1–8. Available from: https://bmcinfectdis.biomedcentral.com/articles/10.1186/s12879-015-1060-3
17. Day SL, Odem-Davis K, Mandaliya KN, Jerome KR, Cook L, Masese LN, et al. Prevalence, Clinical and Virologic Outcomes of Hepatitis B Virus Co-Infection in HIV-1 Positive Kenyan Women on Antiretroviral Therapy. PLoS ONE. 2013 Mar 18;8(3).
18. Ochwoto M, Kimotho JH, Oyugi J, Okoth F, Kioko H, Mining S, et al. Hepatitis B infection is highly prevalent among patients presenting with jaundice in Kenya. BMC Infectious Diseases. 2016 Mar 1;16(1).
19. Muchiri I, Okoth FA, Ngaira J, Tuei S. Seroprevalence of HAV, HBV, HCV, and HEV among acute hepatitis patients at Kenyatta National Hospital in Nairobi, Kenya. East African Medical Journal [Internet]. 2013 Aug 1 [cited 2021 Dec 2];89(6):199–205. Available from: https://www.ajol.info/index.php/eamj/article/view/91508
20. Atina JO, Ogutu EO, Hardison WG, Mumo J. Prevalence of hepatitis A, B, C and human immunodeficiency virus seropositivity among patients with acute icteric hepatitis at the Kenyatta National Hospital, Nairobi. East African Medical Journal [Internet]. 2004 Aug 20 [cited 2021 Nov 9];81(4):183–7. Available from: https://www.ajol.info/index.php/eamj/article/view/9152
21. Wahome E, Ngetsa C, Mwambi J, Gelderblom HC, Manyonyi GO, Micheni M, et al. Hepatitis B Virus Incidence and Risk Factors Among Human Immunodeficiency Virus-1 Negative Men Who Have Sex With Men in Kenya. Open Forum Infectious Diseases. 2017 Jan 1;4(1).
22. Jepkemei KB, Ochwoto M, Swidinsky K, Day J, Gebrebrhan H, McKinnon LR, et al. Characterization of occult hepatitis B in highrisk populations in Kenya. PLoS ONE. 2020 May 1;15(5).
23. Salyani A, Shah J, Adam R, Otieno G, Mbugua E, Shah R. Occult hepatitis B virus infection in a Kenyan cohort of HIV infected anti-retroviral therapy naïve adults. PLoS ONE. 2021 Jan 1;16(1 January).
